# A Systematic Review on COVID-19 Vaccine preferences using Discrete Choice Experiments

**DOI:** 10.1101/2022.06.12.22276299

**Authors:** Adidja Amani, Helen Kamo Selenguai, Yolande Djike Puepi, Iyale Astadjam Dairou, Sebastien Kenmoe, Ariane Nouko, Cheuyem Lekeumo Fabrice Zobel, Suzanne Sap Ngo Um, Paul Olivier Koki Ndombo, Wilfried Mbacham, Pierre Ongolo-Zogo

## Abstract

**Objective:** To determine the attributes of COVID-19 vaccines that influence vaccine acceptance using a DCE through a systematic review.

**Methods:** A systematic search was carried out for articles published up to November 2021 in the PubMed, Psycinfo, Embase, Web of Science, and Global Index Medicus databases. The electronic search algorithm consisted of the terms (Covid-19) AND (Vaccine) AND (discrete choice experiment).

**Findings:** A total of 39 records were retrieved of which 18 duplicates were identified and removed. Of the remaining 21 records, 10 were excluded because they did not use a DCE approach. 11 studies were included in the meta-analyses with a total of 42 795 participants from three WHO regions. We examined 13 attributes of COVID-19 vaccine that influenced acceptance; cost, vaccine efficacy, number of doses, risk of side effects, proof of vaccination, vaccination setting, duration of immunity, doctor’s recommendation, proportion of acquaintances vaccinated, region of vaccine manufacture, background knowledge of herd immunity, life attenuated or mRNA, speed of vaccination development. The four attributes reported to influence COVID-19 vaccine preferences most worldwide were; high vaccine efficacy, low risk of side effects, long duration of immunity and low number of doses of the vaccine.

**Conclusion:** The most preferred COVID-19 vaccine attributes should be taken into account by vaccine manufacturers and public health policy makers for better introduction and acceptance of COVID-19 vaccine to the world.

## Introduction

Coronavirus Disease 2019 (COVID-19) is a pneumonia, caused by a newly identified coronavirus now called Severe acute respiratory syndrome (SARS)-CoV-2 [1]. As of 15 November 2021, SARS-CoV-2 has infected more than 250 million people and caused the deaths of more than 5 million people worldwide, resulting in a case-fatality rate of about 2.1% [2]. Currently, there is no effective treatment for the disease, and the relaxation of non-medical interventions such as the widespread use of face masks, social distancing and home confinement often leads to a re increase in the number of cases [3]. More than a year after the WHO declared COVID-19 a pandemic, it continues to impose a heavy burden on public health systems and affects populations with a negative impact on economies worldwide [4, 5]. Thus, only a vaccine seems to be the right solution to this problem as vaccines are considered the most effective way to control infectious diseases. Since January 2021, several vaccines with efficacy rates of up to 90% have been developed by major laboratories, licensed by the WHO and put on the market to stop the spread of the COVID-19 pandemic [6, 7].

However, the long-term success of the vaccine response to the COVID-19 pandemic will depend on herd immunity, as bringing these vaccines to market is only half the challenge [8]. Thus, herd immunity can be achieved through natural means, stop-and-go strategies, or mass vaccination of the population [9]. However, the path to bringing a new vaccine to market can be politically and economically complicated and COVID-19 vaccines are no exception [10]. In addition, the ideas and opinions of different stakeholders, such as policy makers and health professionals, may affect the uptake of the vaccine to some extent by the general public and may lead to vaccine hesitancy, a major barrier to achieving herd immunity [11].

Vaccine hesitancy can be explained by several factors, including misinformation [1, 12], vaccine novelty and safety, side effects, efficacy, duration of protection, cost, number of doses, route of transmission, location of vaccination sites, disease burden, emergency use authorization, or politicized approval of the vaccination schedule, and this hesitancy may vary considerably from country to country and with the emergence of new SARS-CoV-2 variants [1, 6, 7, 9].

Heterogeneity factors (age, education and financial income) in inter-individual vaccine preferences may have an impact on vaccination [7]. Therefore, there is an urgent need to find out the hindrances to vaccine uptake, which is the most critical factor for the successful adoption of any vaccination program [1]. Understanding the factors that motivate vaccine hesitancy against COVID-19 will highlight targets for public health communication programs that can increase vaccine acceptance.

Although opinion polls have highlighted the issue of COVID-19 vaccine hesitancy, they have been of limited use in preparing mass vaccination campaigns [9]. In contrast, discrete choice experiments (DCEs) have proven to be robust methods for estimating behavior and preference prediction in health economics [13]. These methods allow for accurate estimation of strict refusal and acceptance of vaccination for a range of proposed realistic scenarios [14].

We therefore synthesized data from the DCE literature to highlight the attributes of COVID-19 vaccines that influence vaccine preferences and lead to vaccine hesitancy in order to help public health authorities better introduce COVID-19 vaccination to the general public to achieve herd immunity and stop the pandemic.

## Methods

### Study design and eligibility criteria

The Preferred Reporting Items for Systematic Reviews and Meta-Analyses (PRISMA) approach served as template for this review [15]. The review was declared in the International Prospective Register of Systematic Reviews under number CRD42021265688. All types of published studies evaluating the preferences of Covid-19 vaccine using a discrete choice experiment approach were of interest for this review. Studies from all geographic regions were selected. Review articles were excluded.

### Data sources and search strategy

A systematic search was carried out for articles in English and French published up to November 2021 in the PubMed, Psycinfo, Embase, Web of Science, and Global Index Medicus databases. The electronic search algorithm consisted of the terms that cover (Covid-19) AND (Vaccine) AND (discrete choice experiment). Additional studies were also searched from the reference list of relevant studies.

### Data extraction and synthesis

Duplicate studies from the article search were removed and the remaining articles selected based on a brief overview of the article’s abstract and title. Isolated relevant articles were independently assessed for eligibility and the data extracted by two investigators independently (AA and SK). The information collected covered the first author’s name, year of publication, geographical location, ethnicity, period of the study, participants characteristics (mean age, SD), the total number of study participants, number of adults willing to be vaccinated, inclusion criteria of participants of each study, and the attributes of the vaccine against Covid-19 which influences the acceptance of the vaccine. Discussion and consensus were reached among authors regarding article selection and data extraction.

### Assessment of study quality

A critical appraisal of the quality of the studies included was carried out following the 10 questions for risk of bias assessment as described by Hoy et al. Possible answers to the questions were “yes”, “no”, “unclear” and “not applicable” due to the content of the articles. For all “yes” answers, a score of 1 was assigned and 0 for the others. Articles were considered to be at high, moderate, and low risk of bias when the total score was 0–3, 4–6, and 7–10 respectively. All disagreements were resolved by discussion and consensus between authors.

## Results

### Results of Study Search

A total number of 39 records were identified and 18 duplicates removed (Figure 1). Of the remaining 21 records, 10 were excluded because there were not DCE approach, COVID-19 vaccine or study protocol. This is how for the qualitative review, we conserved 11 records [1, 4, 7, 9, 11, 13, 16, 18, 19, 20, 22].

**Figure 1:**
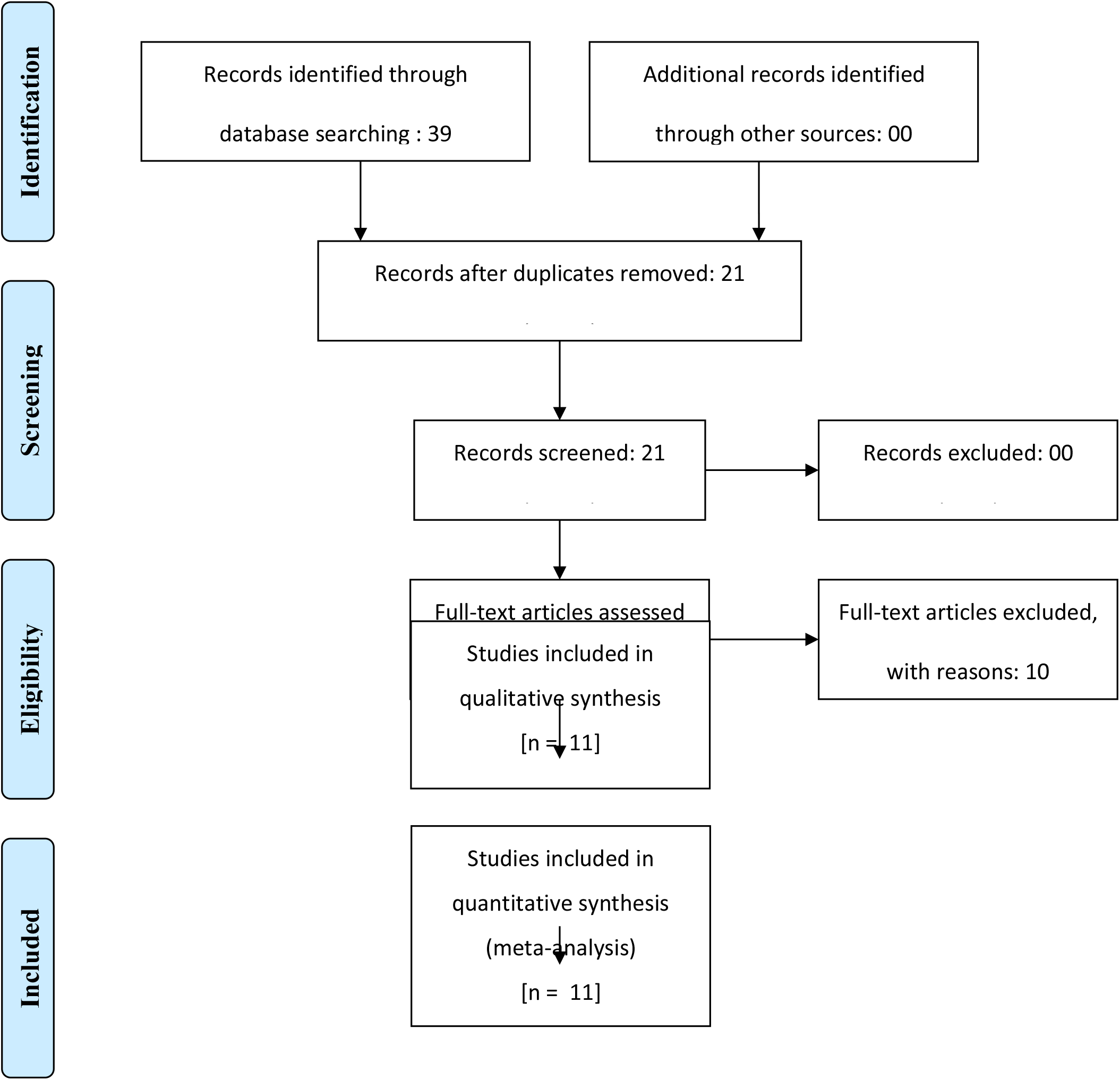
Forest diagram of the study.

### Main findings

Table I summarizes the main characteristics of all studies included in the review and Table II summarizes the attributes investigated in DCE per study and the main findings.

**Table I:**
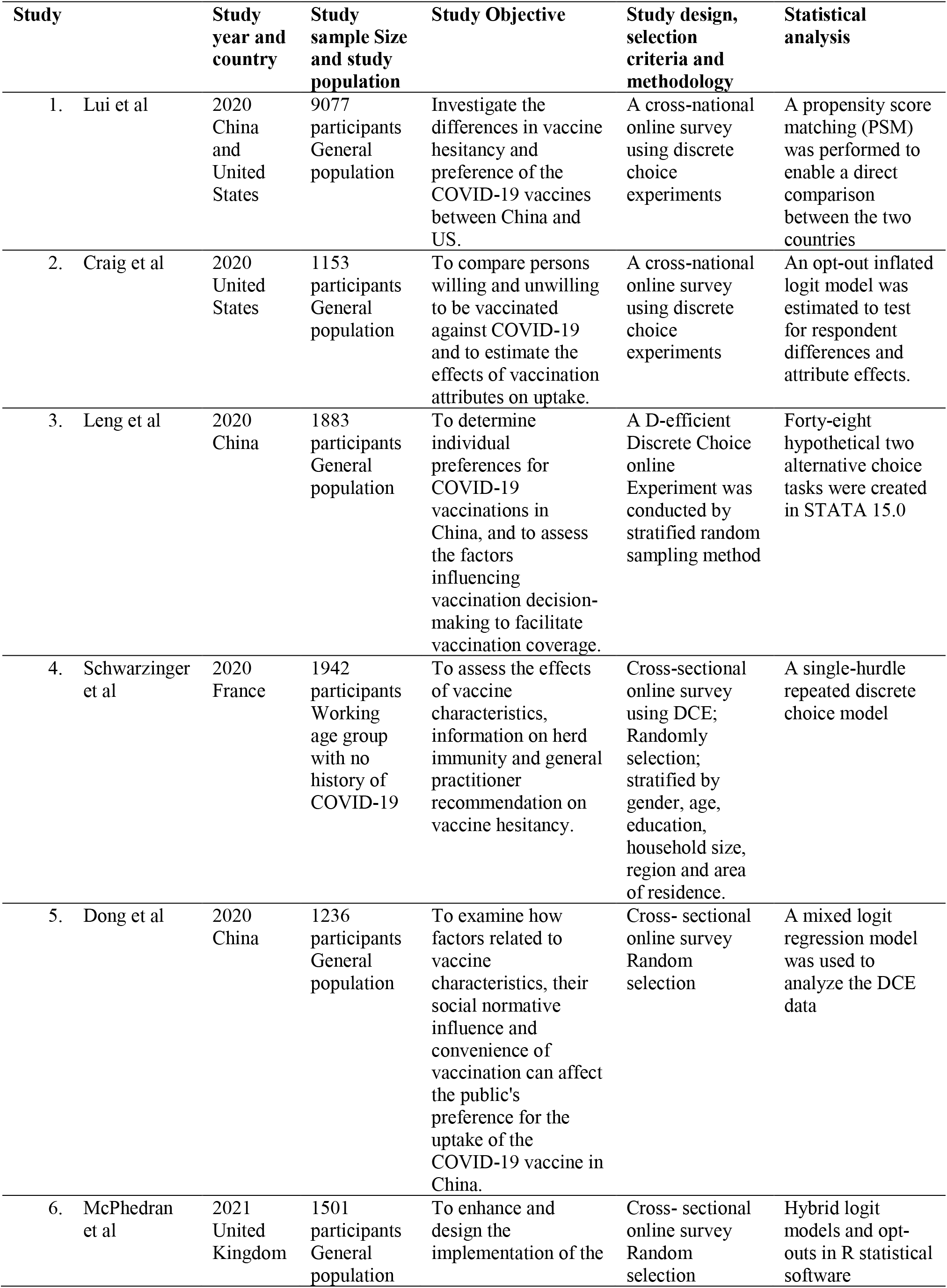
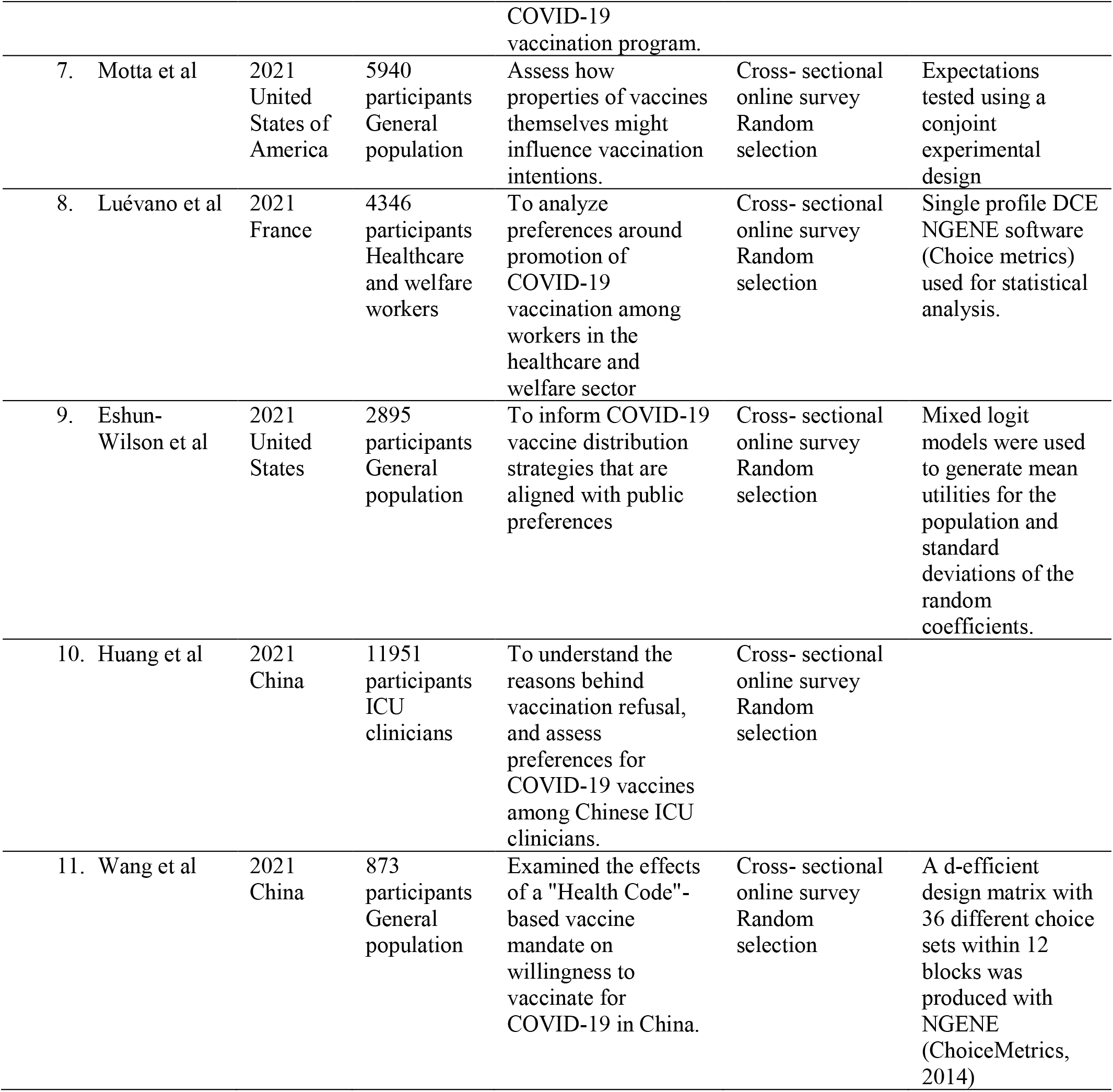
General characteristics of studies included in review

**Table II:** vaccine attributes studied and findings

| Study | Vaccine Attributes investigated using DCE | Main findings |
| --- | --- | --- |
| 1. Lui et al | Cost,<br>Efficacy. | For respondents in China; cost of vaccines was the first factor of hesitancy followed by efficacy of the vaccine and adverse effects of the vaccine.<br>American counterparts; efficacy of the vaccine was the first factor followed by the cost of the vaccine. |
| 2. Craig et al | Proof of vaccination,<br>Vaccination setting,<br>Vaccine effectiveness,<br>Duration of immunity,<br>Risk of severe side effects. | Vaccine effectiveness and safety was the factor with largest effect. |
| 3. Leng et al | Effectiveness,<br>Side effects,<br>Accessibility,<br>Number of doses,<br>Vaccination sites,<br>Duration of vaccine protection,<br>Proportion of acquaintances vaccinated. | Order of vaccine preferences were; vaccine effectiveness, risk of serious side effects, small vaccination fee, fewer doses, larger the protection duration, higher proportion of acquaintances vaccinated. |
| 4. Schwarzing et al | Vaccine efficacy,<br>Risk of serious side effects,<br>Region of vaccine manufacturer,<br>Place of vaccination,<br>Background information about herd immunity,<br>Doctor's advice on vaccination. | Outright refusal was associated with lower perceived severity of COVID-19. Vaccine hesitancy was lower when herd immunity benefits were communicated. Hesitancy increased for vaccines produced in China with 50% efficacy and 1/10 000. |
| 5. Dong et al | Effectiveness (50%, 90%),<br>Protection duration (6 months or 18 months),<br>Adverse effects (1-2days, none),<br>Frequency of injection,<br>Price,<br>Place of origin. | The public preferences in order of highest acceptance; higher effectiveness of vaccine, long protective duration, very few adverse events, being manufactured overseas. |
| 6. McPhedran et al | Level of protection offered,<br>Recommender of the vaccine,<br>Number of doses needed for full protection,<br>Location in which the vaccine is administered,<br>Coverage in the media. | They outlined that efficacy was the factor that most influenced vaccine selection; 90% efficacy preferred 2.8 times more than 70%, and this was more pronounced in individuals above 55 years of age. |
| 7. Motta et al | Country of origin,<br>Effectiveness,<br>Risk of minor side effects,<br>Dose required,<br>Vaccine type,<br>Months spent in development. | They three main factors for vaccine acceptance were the following in order; US made vaccine, followed by a vaccine with 90% effectiveness, and less than 1% risk of minor side effects |
| 8. Luévano et al | Vaccine efficacy,<br>Indirect protection,<br>Vaccine safety,<br>Protection duration,<br>Recommendation/incentive. | They reported high vaccine efficacy (at least 50%) and mRNA vaccine preferred to live attenuated vaccine |
| 9. Eshun-Wilson et al | Vaccination location,<br>Waiting time at vaccination site,<br>Number of doses required,<br>Vaccination enforcement,<br>Who has already received vaccine in community,<br>Vaccine frequency. | Single dose vaccination was preferred to two dose vaccination, vaccination once was preferred to annual vaccination or additional vaccination with boosters, reducing waiting time at vaccination sites was also an important factor. |
| 10. Huang et al | Effectiveness<br>Speed of vaccination development<br>Adverse effects | Vaccine hesitancy among ICU clinicians was due to speed of vaccination development and adverse reactions |
| 11. Wang et al | Price,<br>Origin of vaccine,<br>Probability of fever as side effect,<br>Effectiveness,<br>Location of vaccination,<br>Number of doses, | Willingness to get vaccinated appears to increase if mobile technology prohibits unvaccinated individuals from public spaces. |

The first study retained in our systematic review, used DCE to compare COVID-19 vaccine hesitancy between participants from China and those from America with 5375 and 3702 respondents respectively. For respondents in China; cost of vaccines was the first factor of hesitancy followed by efficacy of the vaccine and adverse effects of the vaccine. While for the American counterparts; efficacy of the vaccine was the first factor followed by the cost of the vaccine. It was also reported that in China; only 82% of study population accepted to be vaccinated, 31.9% of them followed recommendation by a doctor. While in the US; 72.2% of study population accepted to be vaccinated, 50.2% of them following a doctor’s recommendation, 59.4% following advice from friends and family and 64.8% did so freely. China respondents preferred an inactivated vaccine while their American counterparts preferred mRNA vaccine. [1]

The second included article in our qualitative review appraised COVID-19 vaccination preferences in the United States using DCE in 1153 adult participants. They studied the effect of the following attributes on vaccination acceptance; proof of vaccination, vaccination setting, vaccine effectiveness, duration of immunity and risk of severe side effects. Its main findings; less educated people were more likely to be unwilling to take the vaccine (p< 0.001), among those willing, vaccine uptake on the DCE ranged from 61.7-97.75% depending on the vaccine attributes. Vaccine effectiveness and safety was the factor with largest effect. They also reported that offering proof of vaccination and choice of setting for vaccination significantly increased uptake. Duration of immunity had the least influence on vaccine acceptance. [4]

The third study accessed individual preferences for COVID-19 vaccination using DCE in 1883 participants from 6 provinces in China selected by stratifies random sampling methods. The following seven vaccine attributes were used, effectiveness, side effects, accessibility, number of doses, vaccination sites, duration of vaccine protection, proportion of acquaintances vaccinated. They reported that out of the seven, those with highest probability of vaccination were; vaccine effectiveness, risk of serious side effects, small vaccination fee, fewer doses, larger the protection duration, higher proportion of acquaintances vaccinated. They also reported that individuals who were older, lower level of education, lower income, higher trust in vaccine, higher perceived risk of infection displayed a higher probability to be vaccinated. [7]

The fourth study worked on COVID-19 vaccine hesitancy in population in France using DCE, 1942 participants were included out of which 560 refused the vaccine scenario in the DCE. This study examined the following attributes; vaccine efficacy (50%, 80%, 90%, 100%), risk of serious side effects (1 in 10 000, 1 in 100 000), region of vaccine manufacturer (EU, China, USA), place of vaccination (vaccination centre, pharmacy, doctor’s office), background information about herd immunity (no information, information), doctor’s advice on vaccination (no opinion, recommended). Outright refusal and vaccine hesitancy was associated with the female gender, age, lower level of education, poor compliance with vaccines in the past, no report of any chronic disease. Outright refusal was associated with lower perceived severity of COVID-19. Vaccine hesitancy was lower when herd immunity benefits were communicated, hesitancy increased for vaccines produced in China with 50% efficacy and 1/10 000 risk of serious side effects while it decreased with vaccines manufactured in European Union with 90% efficacy and 1/100 000risk of serious side effects. [9]

The fifth study studied public preferences for COVID-19 vaccines in China using DCE. They had 1236 participants, they study the following attributes; effectiveness (50%, 90%), protection duration (6 months or 18 months), adverse effects (1-2days, none), frequency of injection (3 or 2), price (200 CNY or 0), place of origin (imported or domestic). The public preferences in order of highest acceptance; higher effectiveness of vaccine, long protective duration, very few adverse events, being manufactured overseas. It was noted that vaccine price was the least on the list. [11]

The sixth study was done in the UK; they used online DCE to explore preferences of COVID-19 vaccines in 1501 participants. The attributes studied were; level of protection offered (50%, 70%, 90%), recommender of the vaccine (general practitioner (GP), National Health Service (NHS)), number of doses needed for full protection (one or two doses), location in which the vaccine is administered (local GP’s office or mobile vaccination unit), coverage in the media (positive coverage on newspapers, television, radio or positive coverage on WhatsApp, blogs, social media). They outlined that efficacy was the factor that most influenced vaccine selection; 90% efficacy preferred 2.8 times more than 70%, and this was more pronounced in individuals above 55 years of age. For the other attributes; one dose was preferred to 2 doses, recommendation from GP preferred to that of NHS, vaccination at the GP’s office was preferred to vaccination in mobile units, positive coverage on newspapers, television and radio was preferred to that on social media. [13]

The seventh study accessed if COVID-19 vaccine lived up to the American expectations in 5940 participants, they accessed six attributes in the DCE; country of origin (US, UK, China and Russia), effectiveness (50%,70%, 90%), risk of minor side effects (1 in 2, 1 in 10, 1 in 100), dose required (1 or 2), vaccine type (mRNA or weakened virus), months spent in development (9months, 12 months, 15 months). They found out that the three main factors for vaccine acceptance were the following in order; US made vaccine, followed by a vaccine with 90% effectiveness, and less than 1% risk of minor side effects. [16]

The eighth study worked on quantifying healthcare and welfare sector workers’ preferences around COVID-19 vaccination using DCE in 4346 participants in France. They reported high vaccine efficacy (at least 50%) and mRNA vaccine preferred to live attenuated vaccine [18].

The ninth study was on preferences for COVID-19 vaccine distribution strategies in US using DCE with 2895 participants. Single dose vaccination was preferred to two dose vaccination, vaccination once was preferred to annual vaccination or additional vaccination with boosters, reducing waiting time at vaccination sites was also an important factor. They also reported four distinct preference phenotypes; vaccine features, vaccine service delivery, social proof of vaccine safety and indifference to vaccine service delivery [19].

The tenth study examined COVID-19 coverage, concerns, and preferences among Chinese ICU clinicians using DCE. They had 11 951 participants and reported vaccine hesitancy among ICU clinicians was due to speed of vaccination development and adverse reactions [20].

The last study examined if COVID-19 vaccination willingness increase, if mobile technologies prohibit unvaccinated individuals from public spaces using DCE in 873 participants in China. They reported that willingness to vaccine appears to increase if mobile technology prohibits unvaccinated individuals from public spaces and public transportation [22].

## Discussion

The review analysed information from eleven studies from three WHO regions; America (USA), West Pacific (China) and Europe (France and United Kingdom) and four countries; United States of America (3 studies), China (5 studies), United Kingdom (1 study), and France (2 studies). A total of 42, 795 participants from the eleven studies were included in our review. A total of thirteen factors were examined in the DCE from all studies reviewed; cost, vaccine efficacy, number of doses, risk of side effects, proof of vaccination, vaccination setting, duration of immunity provided by the vaccine, doctor’s recommendation, proportion of acquaintances vaccinated, region of vaccine manufacture, background knowledge of herd immunity, if vaccine is life attenuated or mRNA, speed of vaccine development. The main finding of our review is out of the thirteen factors examined in all the studies, there are four factors that were seen to have more influence on COVID-19 preferences than the others; high vaccine efficacy, low risk of side effects, long duration of immunity of the vaccine and reduced number of doses of the vaccine. In addition to these four main preferences mentioned above, participants from China were also concerned about vaccine cost; those from United Kingdom and France were particular on region of vaccine manufacture they preferred vaccines produced in Europe. A peculiarity of participants from the United Kingdom was they preferred receiving the vaccine in a doctor’s office or hospital setting rather than in a mobile vaccination unit. Participants from America also preferred vaccine manufactured in their country in addition to the four main factors for COVID-19 preferences mentioned above.

As seen in Tables 1 and 2; all the studies included in the review using the DCE showing the quality of the review. All the studies used a cross sectional online survey for data collection, with average sample size of 3891participants [873; 11951], nine out of eleven of the studies targeted the general population while two studies targets health care workers, welfare workers and ICU physicians. Selection criteria of participants in all of the studies were randomised sampling method. Statistical methods used in analysis of results mixed and hybrid logit models, opt-out models, propensity score matching in R and Stata. All the studies clearly brought out the vaccine attributes they were investigating in the DCE; on average each study investigated 5 vaccine attributes [2; 7].

Our findings are similar to that reported in systematic review using DCE on parent, provider and vaccine preferences for Human Papilloma vaccine (HPV) which reported high vaccine effectiveness as the attribute that influenced acceptance most. They also reported age of vaccination and cost of vaccine as main preferences for vaccine acceptance[23]. Another systematic review on individual preferences for child and adolescent vaccine attributes reported low risk of side effects, duration of protection and vaccine cost as the most preferred attributes of vaccine. They worked on childhood vaccines, Human Papilloma Vaccine, meningococcal vaccine, Hepatitis B vaccine, diphtheria, tetanus vaccine [24]. A systematic review on choice based experiments on vaccine preferences driving vaccine decision making of different target groups reported vaccine effectiveness, vaccine risk, cost and protection duration as the main factors influencing vaccine decision (Diks et al., 2021). These three systematic reviews have findings similar to our findings although they studied preferences for vaccines other than COVID-19 vaccine; high vaccine effectiveness, low risk of side effects and duration of vaccine protection are reported in all three studies as factors influencing preferences.

On the other hand, a systematic review on effectiveness and acceptability of parental financial incentives and quasi mandatory schemes for increasing uptake of vaccination in preschool children reported that universal parental financial incentives were preferred to quasi-mandatory interventions [26]. Another systematic review on acceptability of parental financial incentives and quasi-mandatory interventions for preschool vaccination reported that incentives and quasi-mandatory vaccination could be effective particularly in disadvantaged groups but were not considered appropriate motivation for vaccination in children [27]. Our study did not examine incentives and quasi-mandatory aspects of COVID-19 vaccination.

The implication of this review for research is that more studies need to be done using DCE in other WHO regions like Africa, Eastern Mediterranean, and South-East Asia in order to know COVID-19 preferences to inform vaccine manufacturers and public health decision makers.

The results from this review should be taken into consideration by public health decision makers for subsequent COVID-19 vaccine introduction. From our findings, countries and continents are encouraged to develop their own vaccine against COVID-19 as most participants are more comfortable taking vaccines manufactured in their countries or continents.

Our review has as limitation the fact that we found very few studies which used the DCE as tool for appraising COVID-19 vaccine preferences limiting the scope of our review. It would have been good to have a study on COVID-19 vaccine preferences in Africa, being the continent, which seems to have the highest hesitancy but we could not find any.

The strength of this review is it covers participants from four continents and over 22 000 people so it gives a good picture of COVID-19 vaccine preferences over the world, it brings to light all the factors for COVID-19 vaccine preferences using DCE. This will inform policy makers on better ways to introduce the Covid-19 vaccine to the World in view of better controlling the pandemic.

## Conclusion

In conclusion our results suggest that vaccine efficacy, risk of side effects of vaccine, duration of immunity provided and the number of doses are the main factors that influence COVID-19 vaccine uptake. This should be taken into consideration by vaccine manufacturers for better introduction of the vaccine throughout the world.

## Data Availability

All data produced in the present work are contained in the manuscript

## Authors’ contributions

(I) Conception and design: A Amani; (II) Administrative support: A Amani; (III) Collection and analysis of data: S Kenmoe, A. Amani; (IV) Data interpretation and discussion: A Nouko, S Kenmoe A. Amani; (V) Manuscript writing: All authors; (VI) Final approval of manuscript: A Amani, S Kenmoe.

